# Tripod transcranial alternating current stimulation at 5-Hz to alleviate anxiety symptoms: a preliminary report

**DOI:** 10.1101/2023.10.17.23296812

**Authors:** Tien-Wen Lee, Chiang-Shan R. Li, Gerald Tramontano

## Abstract

**Introduction:** One of the most common applications of transcranial electrical stimulation (tES) at low current intensity is to induce a relaxed state or reduce anxiety. With technical advancement, different waveforms, montages, and parameters can be incorporated into the treatment regimen. We developed a novel protocol to treat individuals with anxiety disorders by transcranial alternating current stimulation (tACS).

**Methods:** A total of 27 individuals with anxiety disorders underwent tACS treatment for 12 sessions, with each session lasting 25 minutes. tACS at 5 Hz was applied to F4 (1.0 mA), P4 (1.0 mA), and T8 (2.0 mA) EEG lead positions (tripod), with sinewave oscillation between T8 and F4/P4. We evaluated the primary and secondary outcomes using the Beck Anxiety Inventory (BAI) and neuropsychological assessments.

**Results:** Of the 27 patients, 19 (70.4%) experienced a reduction in symptom severity greater than 50%, with an average reduction of BAI 58.5%. All reported side effects were mild, with itching or tingling being the most common complaint. No significant differences were noted in attention, linguistic working memory, visuospatial working memory, or long-term memory in neuropsychological assessments.

**Conclusion:** The results suggest the potential of this novel tripod tACS design as a rapid anxiety alleviator and the importance of a clinical trial to verify its efficacy.

## Introduction

Developed for more than one century and earlier than electroencephalography (EEG), transcranial electrical stimulation (tES) refers to various non-invasive techniques for applying electrical currents to the brain in both research and clinical settings (Bikson et al., 2019). Anxiety reduction has been one of the earliest and most common uses of tES, and many commercial setups, such as Electrosleep and Alpha-Stim, are available for the application of tES (Morriss et al., 2019; Robinovitch, 1914). It was estimated that 3.8 to 25% of people suffered from anxiety disorders (Remes et al., 2016), while less than half received treatment. It is acknowledged that tES is a safe treatment option for many neuropsychiatric conditions (Antal et al., 2017); however, the evidence for its therapeutic effects seems less than compelling (Kavirajan et al., 2014; Shekelle et al., 2018). With the recent technological advancement of tES (Guleyupoglu et al., 2013), we considered here a new protocol to improve the treatment efficacy further.

In Electrosleep, electrical stimulation generally covered the orbital area and mastoid, the delivered waveforms were pulsed at 30–100 Hz, and the intensity was set at the level of a few milliamperes (mA). Electrosleep produces a relaxing effect, which may lead to sleep as an indirect side effect. Electrosleep was thus renamed Cranial Electrostimulation Therapy (Knutson, 1967), and the nomenclature was updated several times over the ensuing decades (Guleyupoglu et al., 2013). NeuroTone 101, which adopted alternating current waveforms, was reviewed, and approved by FDA as the first class-II cranial electrotherapy stimulation (CES) device for treating anxiety, insomnia, and depression (21 Code of Federal Regulations [CFR] 882.5800, 1978). Over time, more CES devices have been marketed for addressing anxiety-related issues. Certain second-generation CES devices moved from transcranial stimulation to applying small square pulsed electric currents through bilateral ear clips with lower frequencies (e.g., 0.5 Hz) and intensity (e.g., hundreds of microamperes) (Bystritsky et al., 2008). These are sometimes referred to as Alpha-Stim because they were believed to improve cortical alpha production, which could contribute to their effectiveness in reducing anxiety.

There are fundamental differences in the designs and stimulation protocols, including the montage, stimulation frequency, and intensity, between Electrosleep and Alpha-Stim. New treatment modalities have been introduced to the treatment of anxiety disorders, including transcranial direct current stimulation (tDCS) and transcranial alternating current stimulation (tACS) (Clancy et al., 2017; Hampstead et al., 2016). The intensity typically falls within the range of several mA, and stimulation sites have been extended to the frontal, parietal, and/or temporal regions to target the frontoparietal network and limbic system. The frontoparietal network plays a critical role in regulating anxiety, and the limbic system is implicated in the manifestation of hyperarousal, worry, apprehension, and re-experiencing of negative emotional states (Engel et al., 2009; Etkin et al., 2006; Hampstead et al., 2016; Phillips et al., 2008). Among the inter-connected limbic area, the amygdala processes fear and threat, and its dysfunction represents a central pathophysiological feature of anxiety (Lee et al., 2013).

Although the application of tACS to the treatment of psychiatric conditions is still in its beginnings, it is of particular interest given that oscillation is a fundamental nature of neural activities (Lee, 2016), and tACS alters neural rhythmicity through the imposition of alternating currents (Ali et al., 2013). With the posited mechanisms of entrainment and cumulative plasticity, tACS may impact the endogenous frequencies (e.g., delta to gamma) of physiological relevance. Clancy et al. applied alpha frequency tACS at 2.0 mA to the occipito-parietal sites and observed a rapid reduction in anxious arousal and aversion to auditory stimuli as well as an enhancement of posterior alpha power that sustained for several days (Clancy et al., 2017). The study highlighted the potential of tACS as a treatment option for anxiety disorders. Alexander et al. applied the alpha tACS over the dorsolateral prefrontal cortex (dlPFC) in patients with major depressive disorder, with sham and 40 Hz stimulation as controls (Alexander et al., 2019). The results showed significantly more responders in the 10 Hz-tACS group; however, the alpha power was reduced in the dlPFC, in contrast to the finding of Clancy et al. (Clancy et al., 2017).

Here, we invented a novel tACS protocol and explored its potential in treating anxiety disorders. In contrast to previous studies, we modulated the frontoparietal network and limbic system simultaneously through F4, P4, and T8 electrode montage according to the 10-10 system of EEG convention. The target dosage was set to 2.0 mA, and the modulating frequency at 5 Hz, as with a transcranial magnetic stimulation study that adopted 5 Hz stimulation for treating comorbid post-traumatic stress disorder (PTSD) and major depression (Philip et al., 2016). The rationale for higher current dosage is based on the observation that the susceptibility of neurons to membrane polarization decreases as the frequency of importing alternating current increases (Anastassiou et al., 2011; Bland and Sale, 2019). To our knowledge, this is the first study that applies tACS at 5 Hz and targets large-scale frontal, parietal, and temporal/limbic regions together in the treatment of anxiety disorders.

## Materials and Methods

### Participants

We conducted a retrospective analysis of patient databanks from our clinics, covering the period between 2018 and 2022. Adult patients with the main diagnoses of generalized anxiety disorder (GAD), social anxiety disorder (SAD), and post-traumatic stress disorder (PTSD), who scored 17 or higher on the Beck Anxiety Inventory (BAI) (Beck et al., 1993), were considered. Patients who met any of the following criteria were excluded: (1) Beck Depression Inventory (BDI) score > 23 (i.e., co-morbid moderate to high depression) (Beck et al., 1987), (2) co-morbid substance use disorders, and (3) somatoform disorders (Kroenke, 2007). A total of 27 patients (16.6 to 74.6 years of age) diagnosed through semi-structured interviews according to the Diagnostic and Statistical Manual of Mental Disorders (DSM-5®) were included in the study (American Psychiatric Association, 2013; First, 2014), who were instructed to continue their current treatments, such as medications, throughout the tES treatment without adjustment of dosage.

Individuals with epilepsy, skull defects, intracranial electrodes, vascular clips or shunts in the brain, cardiac pacemakers, or other implanted biomedical devices, as well as pregnant or lactating conditions, were not eligible for tES in accordance with safety guidelines (Matsumoto, 2017 #12940; Thair, 2017 #13068). Informed written consent was obtained for each clinical patient according to a protocol approved by the Pearl Institutional Review Board (Pearl IRB; https://www.pearlirb.com/).

### Clinical assessments

Before and after treatment, all participants were evaluated by the BAI, BDI, and Personality Assessment Inventory (Beck et al., 1993; Beck et al., 1987; Morey, 1991). A neuropsychological battery covering the domains of attention (test of visual attention; TOVA), linguistic working memory (Letter Fluency), visuospatial working memory (Design Fluency), and long-term memory (free recall) was administered. The working memory tasks were selected from the Delis-Kaplan Executive Function System (D-KEFS) (Delis et al., 2001). Long-term memory was assessed by either Logical Memory II from Wechsler Memory Scale–Fourth Edition (WMS) or California Verbal Learning Test (CVLT) (Delis et al., 2000; Drozdick et al., 2018). The four tasks acted as probes, albeit not thoroughly, to explore the functions of the frontal, parietal, and temporal regions (Fjell et al., 2019; Gron et al., 2001; Marin et al., 2017). After the current intensity reached the optimum (i.e., 2.0 mA), participants filled out a questionnaire about the side effects of tES, including itching/tingling sensation, burning sensation, discomfort or pain, skin lesion, nausea, dizziness, twitching feeling, tinnitus, sleepiness, visual phosphenes, seizure, amongst others.

### tACS administration

We used the FDA-approved Starstim-8 (Neuroelectrics, Inc.; www.neuroelectrics.com) in our clinics. Electric currents were sent to the scalp/brain through electrodes that are connected by wires to a rechargeable battery. Before the electrodes are attached, the scalp needs to be scrubbed with skin preparation gel. The conductive gel is then applied to fill the space between the scalp and electrodes. The above procedures aimed to ensure good contact with the scalp and maintain the impedance level below 5 k ohms (DaSilva et al., 2011).

The montage of electrodes covered the right lateral side of the head at F4, T8, and P4 positions in terms of the 10-10 EEG convention. The peak current intensity was 2.0 milliamperes (mA) for T8 and 1.0 mA for F4 and P4. Alternating sinewave currents oscillated at 5 Hz between T8 and F4/P4. During the neuromodulation session, participants were asked to practice progressive muscle relaxation training by watching a standardized video to enhance the treatment effects (Jacobson, 1938). The setup of tACS is illustrated in Figure 1.

**Fig. 1.** An illustration of the neuromodulation setup. Three electrode positions were wire-connected to a rechargeable battery: F4, P4, and T8.

To help the patients become accustomed to tES, we adopted a dose escalation approach in the first three sessions: 1.0 mA for session 1, 1.5 mA for session 2, and 2.0 mA for session 3. The stimulation lasted 25 minutes, with a 1-minute ramp-up and a 30-second ramp-down phase to reduce skin irritation.

Unlike Alpha-Stim, which can be self-administered at home (Morriss et al., 2019), the Starstim-8 system needs to be operated in clinics. The on-site visits were scheduled for 12 sessions spanning 3 to 4 weeks. Treatment efficacy was evaluated by an index: post-treatment BAI minus pre-treatment BAI)/pre-treatment BAI. Patients with ≥ 50% reduction in symptom severity were regarded as treatment responders.

## Results

All participants tolerated the maximum current at 2.0 mA. The demographic information, diagnoses, and side effect profiles are summarized in Table 1. The symptom severity reduction measured by BAI was 58.5% (SD 16.2%), and the percentage of responders was 70.4%.

**Table 1.**
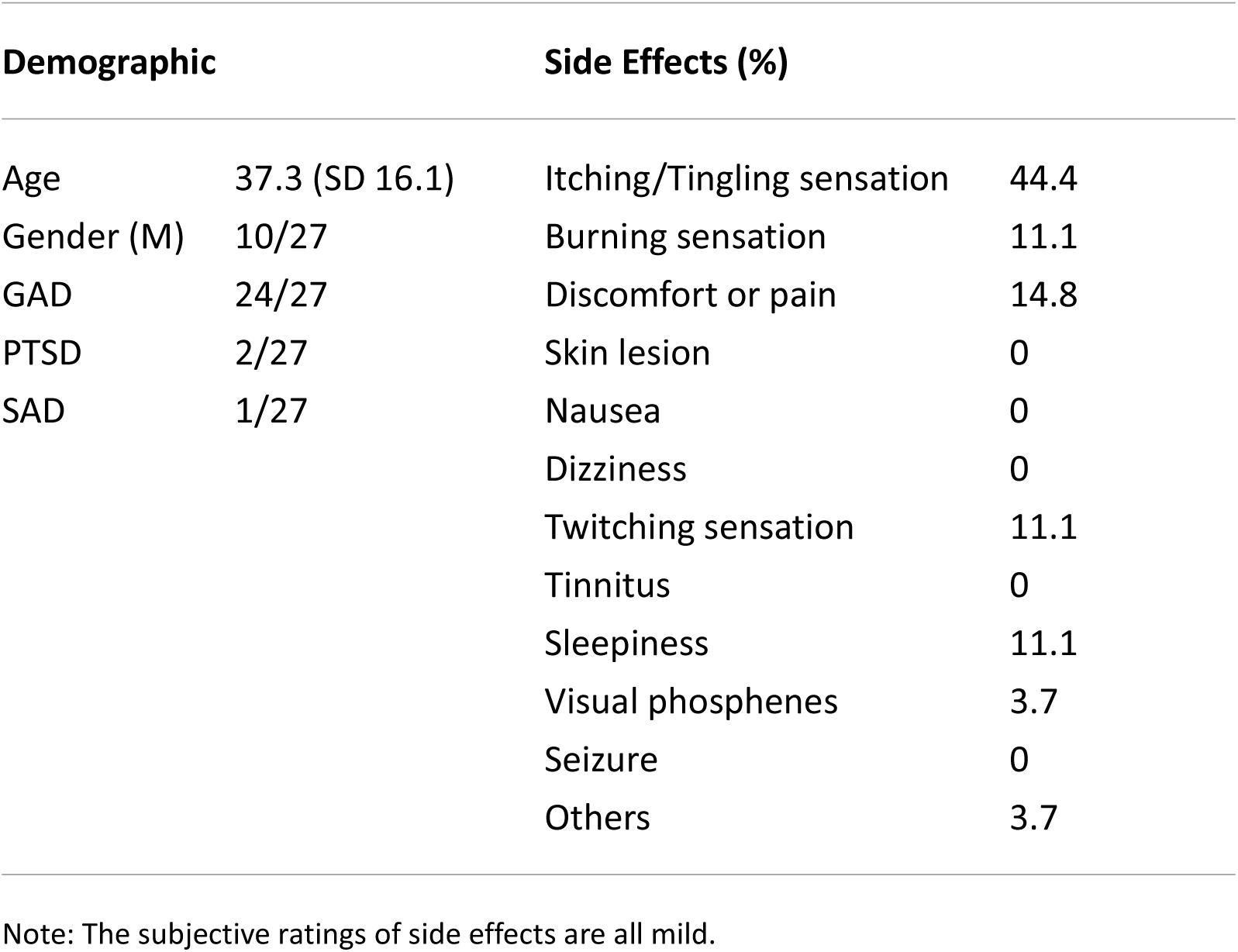
Demographic, diagnosis, treatment session, and side effect profile of tACS at 5 Hz.

Global attention as indexed by the Attention Composite Score of TOVA, working memory as indexed by the total number of correct items in the two fluency tasks of D-KEFS, and long-term memory as indexed by the number of accurate recollections did not show significant differences pre- and post-tACS (all *P* values > 0.05; paired t-tests). The findings are summarized in Table 2.

**Table 2.**
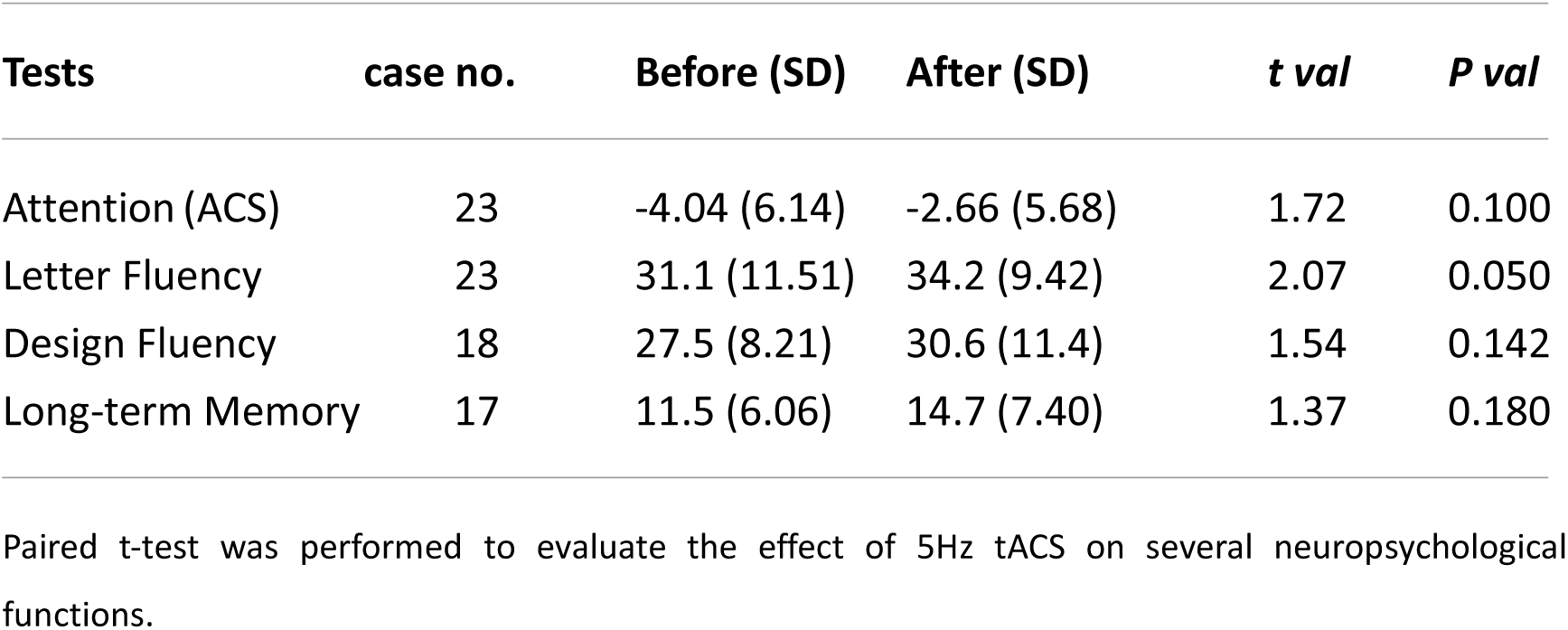
Neuropsychological profile before and after treatment.

| Tests | case no. | Before (SD) | After (SD) | *t val* | *P val* |
| --- | --- | --- | --- | --- | --- |
| Attention (ACS) | 23 | -4.04 (6.14) | -2.66 (5.68) | 1.72 | 0.100 |
| Letter Fluency | 23 | 31.1 (11.51) | 34.2 (9.42) | 2.07 | 0.050 |
| Design Fluency | 18 | 27.5 (8.21) | 30.6 (11.4) | 1.54 | 0.142 |
| Long-term Memory | 17 | 11.5 (6.06) | 14.7 (7.40) | 1.37 | 0.180 |

Paired t-test was performed to evaluate the effect of 5Hz tACS on several neuropsychological functions.

We made a supplementary analysis of five depressive cases whose BDI scores were higher than 23, and the treatment efficacy reflected by BAI reduction was 26.8%, concordant with the impression that patients with co-morbid psychiatric conditions are more resistant to treatment.

## Discussion

The study retrospectively explored the treatment effects of a novel tACS design—5 Hz tACS targeting right frontal, parietal, and temporal regions—for patients with anxiety disorders. The regimen used in this study was within the safety limit of 4 mA and < 60 minutes per day (Antal et al., 2017). The mean symptom severity reduction was 58.5%, and 70.4% patients showed a reduction of > 50%. All reported side effects were mild, and the most frequent complaints were itching or tingling sensation, which was more prominent at the ramp-up phase and attenuated substantially after the current reached its steady plateau. The neuropsychological profile of attention, linguistic working memory, visuospatial working memory, and long-term memory did not show significant differences before and after treatment. Notably, a substantial portion of the patients reported that they experienced remarkable anxiety reduction within just one treatment session, consistent with rapid anxiety relief reported by a previous tACS study (Clancy et al., 2017). Thus, these preliminary results suggest tripod tES at 5 Hz over the right hemisphere as a safe and promising anxiolytic treatment option that may serve as a rapid anxiety alleviator.

Of the anxiolytic tES devices currently on the market, Alpha-Stim and similar CES devices may achieve their efficacy through trigeminal and/or vagus nerve stimulation (via cutaneous vagal afferents) and modulation of the nucleus tractus solitarius, the ventral posteromedial nucleus of the thalamus, and their projections to the forebrain, limbic, and brainstem structures (Adair et al., 2020; Barclay and Barclay, 2014; Butt et al., 2020; Overcash and Kornhauser, 1999). In contrast, our tripod tES design directly targets the frontoparietal and limbic systems. The neuromodulation effect of tES was typically considered confined to the cortical surface. However, recent research has consistently demonstrated the influences of tES on deep cortical and even subcortical regions (Bland and Sale, 2019; Esmaeilpour et al., 2020; Krause et al., 2019; Liu et al., 2018; Louviot et al., 2021). Deep brain stimulation research suggested that modulation of the peri-amygdalar region, including the bed nucleus of the stria terminalis, may reduce anxiety (Luyten, 2020). The core network that supports anxiety is largely subcortical or deep cortical, encompassing the amygdala, insula, ventromedial PFC, orbitofrontal cortex, hippocampus, ventral region of the anterior cingulate cortex, thalamus, hypothalamus, periaqueductal gray, and specific brain stem nuclei (Kolesar et al., 2019), with their downstream projections that regulate behavioral and autonomic manifestations of fear and anxiety (Walker et al., 2003). Incorporating T8 in the tripod design might allow for modulation of the limbic system through the amygdala or the lateral temporal area, which belongs to the default-mode network that conveys limbic information flow (Lee and Xue, 2018). The F4 and P4 electrodes may help to enhance frontoparietal network function and hence the regulation of anxiety (Alexander et al., 2019; Brunyé et al., 2021; Ching et al., 2022; Kavirajan et al., 2014; Lee et al., 2013; Ochsner et al., 2004; Phillips et al., 2008). In support, Ironside et al. found that stimulation of dlPFC by tDCS significantly increased activity in cortical regions associated with attentional control and simultaneously reduced bilateral amygdala reactivity to threatening stimuli (Ironside et al., 2019).

Our treatment design also has practical advantages. Compared with the studies of Alpha-Stim, the treatment duration (session number x session duration) in our treatment was much shorter, but the response rate was comparable. For example, in an earlier Alpha-Stim study, participants received 60 min of daily treatment for six weeks, and the treatment response was approximately 67% (Bystritsky et al., 2008). A relatively large, controlled study with a similar design and parameters showed a treatment efficacy of 54.7% and a response rate of 83% (Barclay and Barclay, 2014). We considered several factors that may account for the effectiveness of treatment with a shorter duration in this innovative approach. First, we excluded patients with BDI higher than 23 and focused on anxiety symptom reduction. Second, the montage of our electrodes allowed for simultaneously modulating the frontoparietal network and limbic system. Third, it is well known that electrical current decreases with traveling distance. Our higher treatment dosage of 2.0 mA may allow the modulation of deep brain structures more effectively. Together, the preliminary results suggest the potential of our tripod tACS and the need for a randomized controlled design and head-to-head statistical comparison of the efficacy with other treatment protocols and experiments. Although the anxiolytic 5-Hz tripod tES seems promising, it may still require a longer treatment duration to solidify and maintain the efficacy, like antidepressant is suggested to continue for several months after symptom improvement.

Anxiety disorders are generally treated with psychotherapy and/or pharmacotherapy. Complete resolution of symptoms may only be seen in approximately 50% of patients (Ballenger, 1999). tES has been regarded as safe (Antal et al., 2017), and hence an ideal adjunct option to potentiate the therapeutic effects of extant treatments. Although emotion processing used to be regarded as mediated by the right hemisphere (Kotz et al., 2006), lateralization is relative, not absolute, especially for anxiety disorders (Gordon et al., 2010). For anxiety patients who do not respond adequately to the right-side 5Hz tripod tES, it may be worthwhile to apply that to the left hemisphere or improvise an alternating left and right treatment sequence to boost the treatment response. It must be cautious that concurrent T7 and T8 tACS might enhance the bi-hemispheric limbic interaction and worsen the psychiatric conditions. Nevertheless, our unreported cases showed that anti-phase tACS over T7 and T8 simultaneously could reduce limbic crosstalk and is another potential anxiolytic design. Like the trend of “combo” in psychopharmacology for refractory cases, a combination of different variants of 5Hz tACS may be worthy of consideration as a potentiation strategy for non-invasive neuromodulation protocol.

In particular, less than half of those suffering from anxiety disorders are estimated to receive treatment (Alonso et al., 2018). Home-based tES devices are now more accessible and user-friendly than ever. With clinicians able to adjust the parameters remotely, this technology becoming highly feasible for more patients and a broader range of social scenarios (e.g., in a pandemic when medical service is severely compromised).

### Conclusion

Despite the vast and high enthusiasm for tES, only a few studies have explored the therapeutic effects of tACS for anxiety. Based on the understanding of brain network dysfunction in the pathophysiology of anxiety disorders, we developed a tripod 5 Hz tACS protocol and demonstrated its rapid effects on reducing anxiety symptom severity. The side effects were minimal. Double-blind and sham-controlled research is warranted to verify its efficacy and role as an adjunct treatment for anxiety disorders.

## Authors Contributions

All authors contributed intellectually to this work. TW Lee carried out the analysis and wrote the first draft. All authors revised and approved the final version of the manuscript.

## Data Availability

All data produced in the present study are available upon reasonable request to the authors

## Acknowledgments

This work was supported by NeuroCognitive Institute (NCI) and NCI Clinical Research Foundation Inc. We would like to thank Almeida Sergio for helping prepare the research material.

## Statements and Declarations

All authors declare no conflicts of interest.

## Compliance with ethical standards

This research analyzed the databank collected from 2018 to 2022. The authors assert that all procedures contributing to this work comply with the ethical standards of the relevant national and institutional committees on human experimentation and with the Helsinki Declaration of 1975, as revised in 2008.

